# Serum metabolic signatures are associated with anti-drug antibody development in rheumatoid arthritis patients treated with adalimumab

**DOI:** 10.1101/2025.08.22.25334214

**Authors:** Tanya Pretorius, Alexandra E Oppong, ABIRISK consortium, Pierre Dönnes, Jessica J Manson, Elizabeth C Jury

## Abstract

**Objectives:** Development of anti-drug antibodies (ADAs) is a barrier to long-term efficacy of biologic therapies in rheumatoid arthritis (RA), but no biomarkers exist to predict ADA formation. This study explored the potential of serum metabolomics to predict development of ADAs to adalimumab in patients with RA.

**Methods:** Serum from patients with RA (n=47), treatment naïve for tumour necrosis factor-alpha inhibitor therapy, were collected before, Month(M)1 and M12 following initiation of adalimumab therapy as standard of care. Sera were tested for ADAs and patients were stratified according to M12 ADA status (ADA-positive n=21; ADA-negative n=26). Serum metabolomics was performed using a NMR-based platform. Metabolomic and clinical data were analysed using machine learning (ML) to develop a signature associated with ADA development.

**Results:** ML analysis of baseline serum metabolomics and clinical data identified a signature that distinguished patients according to their future M12 ADA status (ADA-positive/ADA-negative) prior to first adalimumab treatment (area under the receiver operator curve, AUC-ROC=0.78), which out-performed clinical parameters alone (AUC-ROC=0.78). Metabolites related to cholesterol transport including large high and very low-density lipoproteins (L-HDL/VLDL) and small low density-lipoprotein (S-LDL) and clinical markers body mass index (BMI) and erythrocyte sedimentation rate were top discriminating features. Patients stratified as ADA-positive/ADA-negative at baseline also had different serum metabolic responses to adalimumab at M1 and M12. Finally, a putative predictive score for future ADA status was generated comprising L-HDL, L-LDL, extra-large VLDL subsets and BMI.

**Conclusion:** These results support the potential of serum metabolomics as a predictive tool for immunogenicity risk in RA.

**Graphical abstract:** 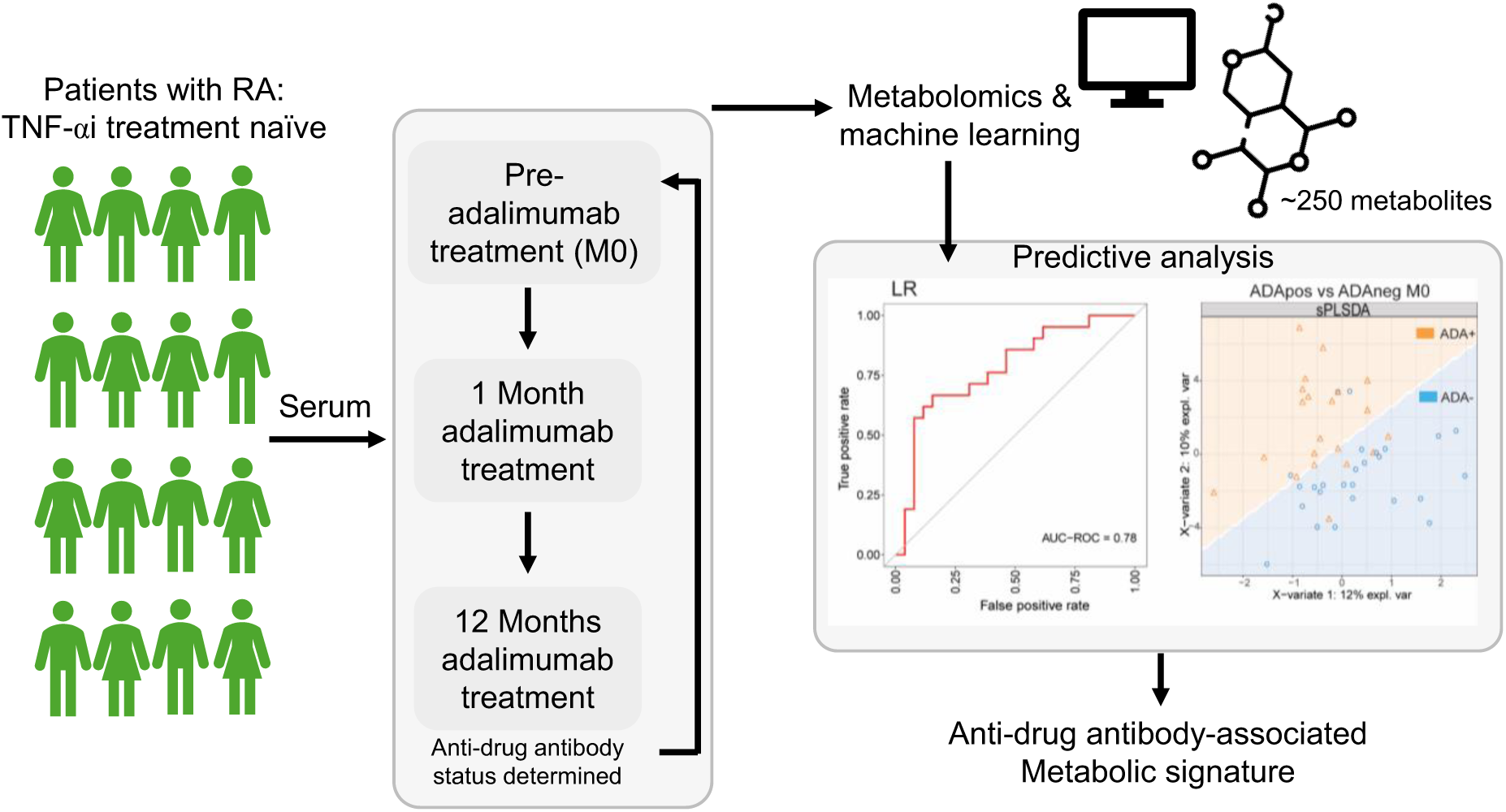

**Key messages:**

- Machine learning models identified serum metabolomic signatures associated with future treatment immunogenicity.
- Lipid-related metabolites suggest changes in lipid metabolism could influence ADA susceptibility.

## Introduction

Rheumatoid arthritis (RA) is a chronic autoimmune disease characterized by inflammation of the joints, where the immune system mistakenly attacks joint tissue(1, 2). While the exact pathophysiology remains unclear, autoantibodies and reactive immune cells play a key role and are common targets for therapeutic interventions(1–3). Current treatments aim to slow disease progression and prevent joint damage through immunosuppression, modulation of immune signaling, and antioxidant effects(4). Biologic disease-modifying anti-rheumatic drugs, such as tumour necrosis factor-alpha (TNF⍺) inhibitors, are often prescribed long-term to specifically target immune components(4, 5).

Despite these advancements, many patients discontinue treatment within two years due to adverse effects or perceived lack of effectiveness(6, 7). A major cause of this reduced efficacy is the development of anti-drug antibodies (ADAs), which can neutralize the therapeutic effects of biologics, may induce immune-mediated adverse reactions and increase healthcare costs(8, 9). In patients with RA treated with TNF⍺ inhibitors, 30-50% develop ADAs within the first year, significantly lowering serum drug levels(10), making this class of drugs a major focus of immunogenicity studies. ADA formation is influenced by factors such as protein structure, dosage, concomitant medications, and immune status(8). Although humanized monoclonal antibodies have reduced immunogenicity, they still carry a risk of anti-idiotype responses, especially when administered subcutaneously, as seen with adalimumab(11, 12). Given the impact of ADAs on patient outcomes, ADA detection and prediction have become crucial areas of research.

Metabolomics offers a promising approach to understanding immunogenicity in RA. Metabolites, byproducts of biochemical processes, reflect the current immune state and can reveal underlying mechanisms driving undesirable immune responses(13). This approach has been valuable in understanding inflammation in diseases like inflammatory bowel disease and COVID-19, and in autoimmune conditions such as RA, multiple sclerosis, and lupus(14–18).

Recent studies have explored the potential of artificial intelligence to analyse metabolomic data and predict ADA formation. In multiple sclerosis patients treated with interferon-β, predictive models using pre-treatment metabolomic data were able to forecast ADA development(16). The Anti-Biopharmaceutical Immunization: Prediction and Analysis of clinical relevance to minimise the RISK (ABIRISK) consortium has applied machine learning (ML) across various autoimmune diseases to assess drug immunogenicity, finding common immune responses across diseases and treatments(19). Identifying these shared mechanisms may allow for more effective, generalized strategies to prevent ADA formation.

This study focuses on determining whether serum metabolomic signatures can predict immunogenicity to TNF⍺-inhibitor treatments in patients with RA and identify metabolites associated with ADA development. The aims are to identify metabolites linked to ADA development in patients with RA and generate an early predictive signature for ADA development in patients with RA.

## Methods

### Patient cohort

Patients with RA (n=47) were recruited as part of the Anti-Biopharmaceutical Immunization: Prediction and Analysis of Clinical Relevance to Minimize the Risk of Immunization in Rheumatoid Arthritis Patients (ABI-RISK) consortium, which is registered on the ClinicalTrials.gov website (NCT02116504). This was a prospective study of patients with RA from four countries (France, Italy, the Netherlands, and the UK). Eligible patients were 18 years or older, had a diagnosis of RA, and were initiating a new anti-TNF inhibitor (adalimumab). Patients were treated according to the local recommendation of their country and were followed for up to 18 months. Recruitment spanned from March 3, 2014, to June 21, 2016. Ethical approval was granted by the Comité de Protection des Personnes Ile de France VII for France (reference 13–048); the Medical Ethical Committee of the Academisch Medisch Centrum, Amsterdam for the Netherlands (reference 2013–304#B20131074); the Local Ethics Committee of Azienda Ospedaliero Universitaria Careggi for Italy (reference 2012/0035982); and the National Research Ethics Service Committee London, City and East (reference 14/LO/0506) for the UK. All participants provided informed written consent.

Serum samples were collected at baseline (M0) prior to first treatment with adalimumab and one month (M1) and 12 months (M12; two individuals did not have data at M12 so ADA status at M15-18 was used) after adalimumab treatment initiation. Demographic, treatment and clinical data, including sex, age, ethnicity, BMI, smoking status, disease activity score (DAS), liver/kidney function markers (aspartate aminotransferase; AST, alanine aminotransferase; ALT, creatinine; CREAT), inflammation markers (C reactive protein; CRP, erythrocyte sedimentation rate; ESR), and immune cell measures (whole blood count; WBC, neutrophil count; NEUT, and lymphocyte count; LYM), were recorded. Smoking status was categorized as ‘never,’ ‘current,’ or ‘quit.’ Serum sampling and consequent predictive analysis were carried out as summarised in **Figure 1A-B**.

**Figure 1:**
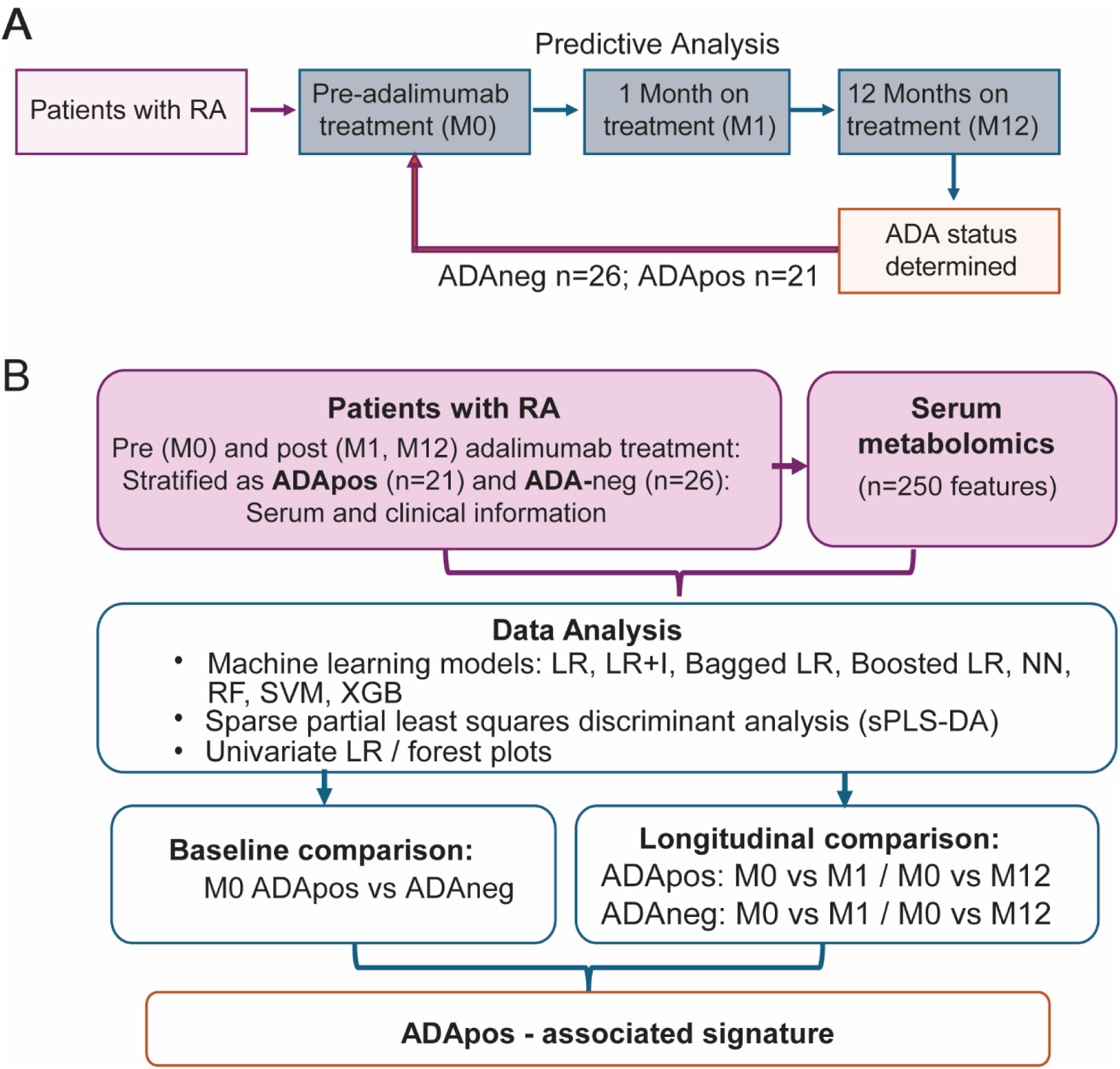
Stratification process and data analysis workflow. **(A)** Patients diagnosed with RA were recruited and treated as standard of care by their treating clinician. Patient and clinical data and biological samples were collected at baseline (month, M0), M1 and M12 following treatment initiation. Serum was tested for binding anti-drug antibodies (ADA). ADA positivity was defined as ADApositive (pos) if titres were >18 ng/mL at least once within 12 months of starting treatment according to the ABIRISK guideline (20). **(B)** Study plan of data analysis. Predictive analysis was conducted using data from all timepoints. Clinical and metabolomic data were processed for analysis using imputation, homology reduction, scaling (mean) and centring (standard deviation) (see methods). Metabolites that ranked among the top ten in more than one machine learning (ML) model or appeared in both sparse partial least square discriminant analysis and univariate LR analyses were analysed using AutoScore (21) to develop an ADApos-associated signature. Abbreviations: LR, logistic regression; LR=I, logistic regression plus interactions; NN, neural network; RF, random forest; SVM, support vector machine; XGB, extreme gradient boosting.

### Sample analysis

#### Anti-drug antibody (ADA) assessment

Binding ADAs were quantified using electrochemiluminescence using the Meso Scale Discovery platform (MSD; Meso Scale Diagnostics LLC), which was performed by the Inflammation, Microbiome and Immunosurveillance Laboratory (Institut National de la Santé et de la Recherche Médicale UMR 996), as previously described (19). Patients were stratified as ADA-negative (ADAneg) or ADA-positive (ADApos). Patients were defined as ADApos if relative electrochemiluminescence (RECL) >2,08 and percentage inhibition >23,8% measured at least once within 12-18 months of starting treatment (**Figure 1A**).

#### Metabolomics

Serum metabolites (n=250) were measured using an established nuclear magnetic resonance (NMR)-spectroscopy platform (Nightingale Health)(22). These included absolute concentrations, ratios, and percentages of lipoprotein: apolipoproteins (Apo), (very) low density ((V)LDL), intermediate density (IDL) and high density (HDL) lipoprotein particles (P) of different sizes ranging from chylomicrons and extremely large (XXL), very large (XL), large (L), medium (M), small (S), and very small (XS); lipoprotein composition including cholesterol (C), cholesterol esters (CE), free cholesterol (FC), triglycerides (TG), phospholipids (PL), total lipid (L); and glycoprotein A (GlycA), amino acids, glycolysis and fluid balance metabolites (See **Supplementary Table S1** for list of metabolites and abbreviations used).

### Data processing

#### Missing data

No features had more than 10% missingness. Features with <10% missing data were imputed using k-nearest neighbours (k=5) using bnstruct in R (23).

#### Homology reduction

To account for biological interactions, features with a correlation coefficient of more than 0.95 were examined and the feature with the greatest mean absolute correlation removed (caret(24), R). Following homology reduction, the number of measures remaining for each comparison were n=131 (ADApos-M0 vs ADAneg-M0), n=126 (ADAneg-M0 vs ADAneg-M1), n=127 (ADAneg-M0 vs ADAneg-M12), n=135 (ADApos-M0 vs ADApos-M1), and n=134 (ADApos-M0 vs ADApos-M12).

#### Data optimisation

Preliminary analysis tested the best combination of demographic and metabolomic data to use in the ML analysis. For all comparisons, reduced datasets worsened performance, except the baseline ADApos-M0 vs ADAneg-M0 comparison, wherein using percentage-only metabolite measures with a reduced number of clinical markers performed best. For all other ML models, the input dataset was the total metabolites after homology reduction and all clinical markers.

### Predictive models

Eight supervised learning algorithms were implemented: Logistic regression (LR), LR with interactions (LR+I), bagged LR, boosted LR, neural network (NN), random forest (RF), support vector machine (SVM), and eXtreme Gradient Boosting (XGBoost). Models were generated from metabolite features at M0, M1 and M12 and clinical features at M0. Parameters were tuned with the “train” function of caret(24) and are summarised in **Supplementary Table S2**.

The comparisons examined included: (i) ADApos vs ADAneg at baseline (M0)(ii) M1 vs M1 and M0 vs M12 in patients stratified as ADApos; (iii) M1 vs M1 and M0 vs M12 in patients stratified as ADAneg (**Figure 1B**). Measurements were centred on the mean and scaled to the standard deviation using matrixStats(R)(25). Caret(24) was utilised for model training in all supervised learning algorithms.

#### Model performance

Ten-fold cross-validation was used to prevent overfitting and to evaluate model performance. Specificity and sensitivity performance metrics were calculated from the confusion matrices. The receiver operating characteristic area under curve (AUC-ROC) was also assessed.

### Data visualisation and statistical testing

Outputs from ML models, sparse Partial Least Squares Discriminate analysis (sPLS-DA), and univariate LR identified metabolic signatures for ADA prediction. RStudio (v4.2.3) was used for all analyses.

#### Univariate logistic regression/forest plots

Data analysed with univariate LR were adjusted for age, sex, ethnicity, country, and smoking status. Results were plotted as forest plots using vignettes of “ggforestplot” (v0.1.0, R, Nightingale Health). The differences between the subgroups were quantified as logarithm of odds ratios.

#### T-tests & Youden index

T-tests were conducted using the Biomarker Analysis feature of MetaboAnalyst(26) (v6.0) to quantify the log2(fold change) and associated p-values. The average values of each metabolite were tested for statistically significant differences between sample groups (p<0.05). Data were not transformed nor scaled prior to analysis. Box plots were created with MetaboAnalyst (26), providing the Youden index and associated sensitivities and specificities for each metabolite.

#### Sparse Partial Least Squares Discriminate Analysis

sPLS-DA is a supervised clustering ML approach that combines classification and parameter selection. sPLS-DA was performed using the mixOmics R package (v6.22.0) (27), with parameters optimised to achieve the lowest overall error rate, with ten-fold cross validation. Variable loading plots presented the top features selected to discriminate between component one and two. sPLS-DA results were visualised using ggplot2 (v3.5.1, R)(28).

### Assessing metabolite significance

Metabolites associated with ADA-positivity were identified using outputs from ML models, sPLS-DA, and univariate analyses. Metabolites that ranked among the top ten in more than one ML model or appeared in both sPLS-DA and univariate logistic regression analyses were included in a shortlist for each comparison. These metabolite sets were then assessed using AutoScore(21) (v.1.0.0, R), a ML-based tool for generating clinical scoring models.

*Model generation:* AutoScore (21) operates in several modules—variable ranking, transformation, score derivation, model selection, fine-tuning, and evaluation. The baseline dataset (ADAposM0 vs. ADAnegM0) was split into training (70%) and testing (30%) sets. RF ranked variables, and multivariable LR generated the final point-based scoring system. The model’s performance was assessed using AUC-ROC and fine-tuned based on the Youden Index. The final model, derived from the M0 vs M1 shortlist was evaluated for its ability to distinguish ADApos from ADAneg patients to identify metabolites associated with ADA predisposition.

## Results

### Metabolite profiles in ADApos patients differ from ADAneg patients at M0, M1 and M12

Patients with RA, who were TNF-inhibitor treatment naïve, were stratified as ADApos (n=21) or ADAneg (n=26) based on their ADA status at M12 after initiation of adalimumab therapy (**Figure 1A**). Most patients developed ADAs in the first month following treatment commencement (**Supplementary Table S3**). There were no significant differences in baseline (M0) demographic, routine clinical assessments and treatment between patients who remained ADAneg and those that developed ADA over the course of the study (**Table 1**). Furthermore, applying ML models to clinical parameters alone could not accurately classify ADApos and ADAneg patients (AUC-ROCs between 0.38-0.66) (**Supplementary Table S4)**. However, combining clinical features with serum metabolite markers was able to improve prediction of ADA development in these patients. Three machine learning models distinguished ADApos from ADAneg patients at M0 (prior to first treatment with adalimumab) with an AUC-ROC >0.7 (**Figure 2A**). Metabolites related to cholesterol transport (L-HDL-FC%, L-VLDL-TG%, Omega-3%, and S-LDL-FC%) were important for every model, while demographic and clinical markers, including BMI, ESR, smoking status, and sex, appeared in two of the models, together with other cholesterol-related metabolites including IDL-FC% and XXL-VLDL-C% (**Figure 2A**). The top performing LR model was able to classify 76.5% of patients correctly with an AUC-ROC of 0.78 (**Figure 2B**). The sPLS-DA model also clustered patients correctly (**Figure 2C)** with the top features driving clustering also identified in the ML models (L-HDL-FC%, S-LDL-FC% and XXL-VLDL-C%). Combined loadings from components 1 and 2 stratified patients with an AUC-ROC=0.882 (**Supplementary Figure S1A-C)**. However, there were no metabolomic differences between ADApos and ADAneg patients at M0, using univariate LR correcting for demographic/clinical features.

**Figure 2:**
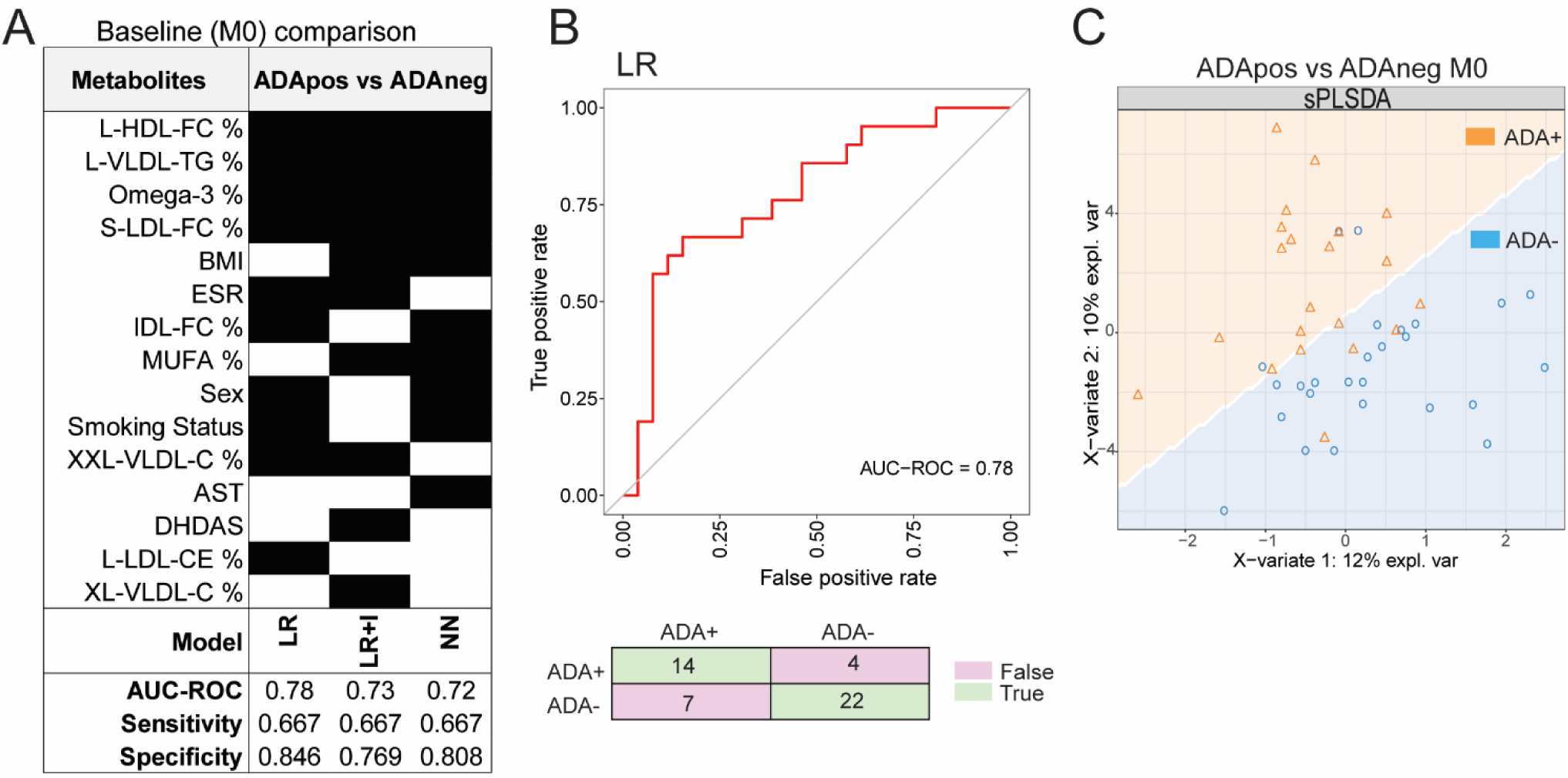
Baseline metabolites combined with clinical features classified patients based on their ADA status determined at M12 following treatment initiation. Patients with RA prior to first treatment with adalimumab were stratified according to the anti-drug antibody (ADA) status at M12 after treatment initiation (ADApos, n=21; ADAneg, n=26). **(A)** M0 clinical data and serum metabolites were analysed using multiple machine learning models (see Methods). Summary of the top features identified by machine learning models with AUC-ROC >0.7 are shown. **(B)** Receiver operator characteristic (ROC) curve and confusion matrix of the top performing logistic regression (LR) model in (A). **(C)** M0 metabolite and clinical data was also analysed using a sparse partial least square discriminant analysis model (see Supplementary Figure 1A-C for ROC and loadings for components 1 and 2).

**Table 1:**
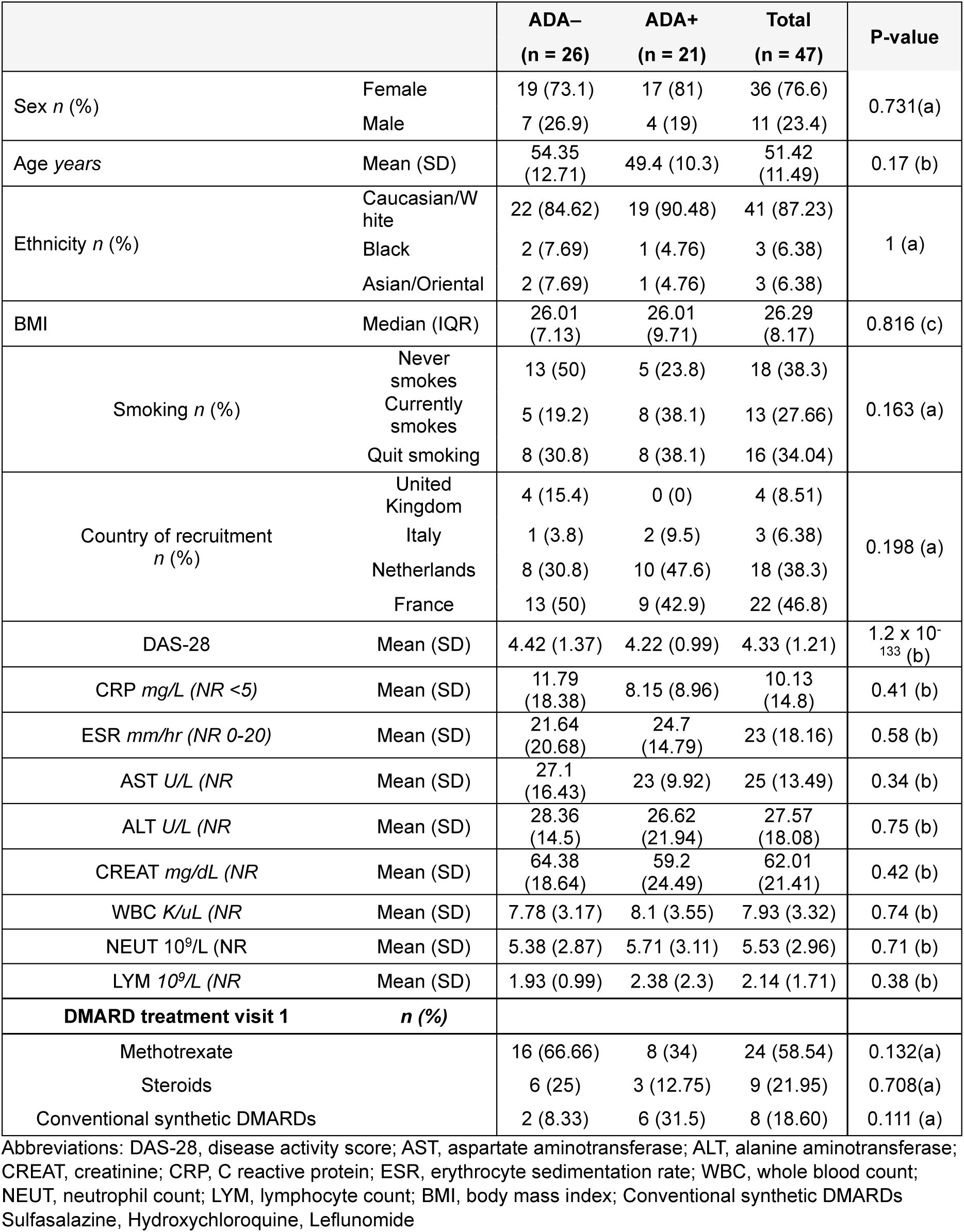
Cohort characteristics. Demographic characteristics and clinical markers were compared between ADApos and ADAneg patients. Statistical comparisons were made using (a) Fisher’s exact test, (b) pairwise comparisons using T-tests with pooled standard deviation, or (c) Mann-Whitney U Test (Wilcoxon Rank-Sum Test). BMI, body mass index; IQR, interquartile range.

|  |  | **ADA–**<br>**(n = 26)** | **ADA+**<br>**(n = 21)** | **Total**<br>**(n = 47)** | **P-value** |
| --- | --- | --- | --- | --- | --- |
| Sex *n* (%) | Female | 19 (73.1) | 17 (81) | 36 (76.6) | 0.731(a) |
|  | Male | 7 (26.9) | 4 (19) | 11 (23.4) |  |
| Age *years* | Mean (SD) | 54.35<br>(12.71) | 49.4 (10.3) | 51.42<br>(11.49) | 0.17 (b) |
| Ethnicity *n* (%) | Caucasian/W<br>hite | 22 (84.62) | 19 (90.48) | 41 (87.23) | 1 (a) |
|  | Black | 2 (7.69) | 1 (4.76) | 3 (6.38) |  |
|  | Asian/Oriental | 2 (7.69) | 1 (4.76) | 3 (6.38) |  |
| BMI | Median (IQR) | 26.01<br>(7.13) | 26.01<br>(9.71) | 26.29<br>(8.17) | 0.816 (c) |
| Smoking *n* (%) | Never<br>smokes | 13 (50) | 5 (23.8) | 18 (38.3) | 0.163 (a) |
|  | Currently<br>smokes | 5 (19.2) | 8 (38.1) | 13 (27.66) |  |
|  | Quit smoking | 8 (30.8) | 8 (38.1) | 16 (34.04) |  |
| Country of recruitment<br>*n* (%) | United<br>Kingdom | 4 (15.4) | 0 (0) | 4 (8.51) | 0.198 (a) |
|  | Italy | 1 (3.8) | 2 (9.5) | 3 (6.38) |  |
|  | Netherlands | 8 (30.8) | 10 (47.6) | 18 (38.3) |  |
|  | France | 13 (50) | 9 (42.9) | 22 (46.8) |  |
| DAS-28 | Mean (SD) | 4.42 (1.37) | 4.22 (0.99) | 4.33 (1.21) | 1.2 x 10<sup>-133</sup> (b) |
| CRP *mg/L* (NR <5) | Mean (SD) | 11.79<br>(18.38) | 8.15 (8.96) | 10.13<br>(14.8) | 0.41 (b) |
| ESR *mm/hr* (NR 0-20) | Mean (SD) | 21.64<br>(20.68) | 24.7<br>(14.79) | 23 (18.16) | 0.58 (b) |
| AST *U/L* (NR | Mean (SD) | 27.1<br>(16.43) | 23 (9.92) | 25 (13.49) | 0.34 (b) |
| ALT *U/L* (NR | Mean (SD) | 28.36<br>(14.5) | 26.62<br>(21.94) | 27.57<br>(18.08) | 0.75 (b) |
| CREAT *mg/dL* (NR | Mean (SD) | 64.38<br>(18.64) | 59.2<br>(24.49) | 62.01<br>(21.41) | 0.42 (b) |
| WBC *K/uL* (NR | Mean (SD) | 7.78 (3.17) | 8.1 (3.55) | 7.93 (3.32) | 0.74 (b) |
| NEUT 10<sup>9</sup>/L (NR | Mean (SD) | 5.38 (2.87) | 5.71 (3.11) | 5.53 (2.96) | 0.71 (b) |
| LYM 10<sup>9</sup>/L (NR | Mean (SD) | 1.93 (0.99) | 2.38 (2.3) | 2.14 (1.71) | 0.38 (b) |
| **DMARD treatment visit 1**<br>***n* (%)** |  |  |  |  |  |
| Methotrexate |  | 16 (66.66) | 8 (34) | 24 (58.54) | 0.132(a) |
| Steroids |  | 6 (25) | 3 (12.75) | 9 (21.95) | 0.708(a) |
| Conventional synthetic DMARDs |  | 2 (8.33) | 6 (31.5) | 8 (18.60) | 0.111 (a) |

Abbreviations: DAS-28, disease activity score; AST, aspartate aminotransferase; ALT, alanine aminotransferase; CREAT, creatinine; CRP, C reactive protein; ESR, erythrocyte sedimentation rate; WBC, whole blood count; NEUT, neutrophil count; LYM, lymphocyte count; BMI, body mass index; Conventional synthetic DMARDs Sulfasalazine, Hydroxychloroquine, Leflunomide

### Patients stratified as ADApos and ADAneg had altered metabolic responses to treatment at M1 and M12

Next, we assessed the response to therapy in patients stratified as either ADApos or ADAneg comparing M0 vs M1 and M0 vs M12+ within each group. Several models identified differences (AUC-ROC >0.7) in metabolite and clinical features over the time course of the study (**Figure 3A-C, Supplementary Fig S2A-D** for top performing model of each comparison). Not surprisingly, DAS-28 was an important feature for classifying both ADApos and ADAneg patients in all comparisons, highlighting its role in assessing response to treatment in RA (**Supplementary Fig S2E)**. Acetoacetate and acetate were also important features showing reduced and increased abundance respectively at M1 and M12 vs baseline supporting an ADA-independent role in response to treatment with adalimumab (**Figure 3A-C, Supplementary Fig S2E-G**).

**Figure 3:**
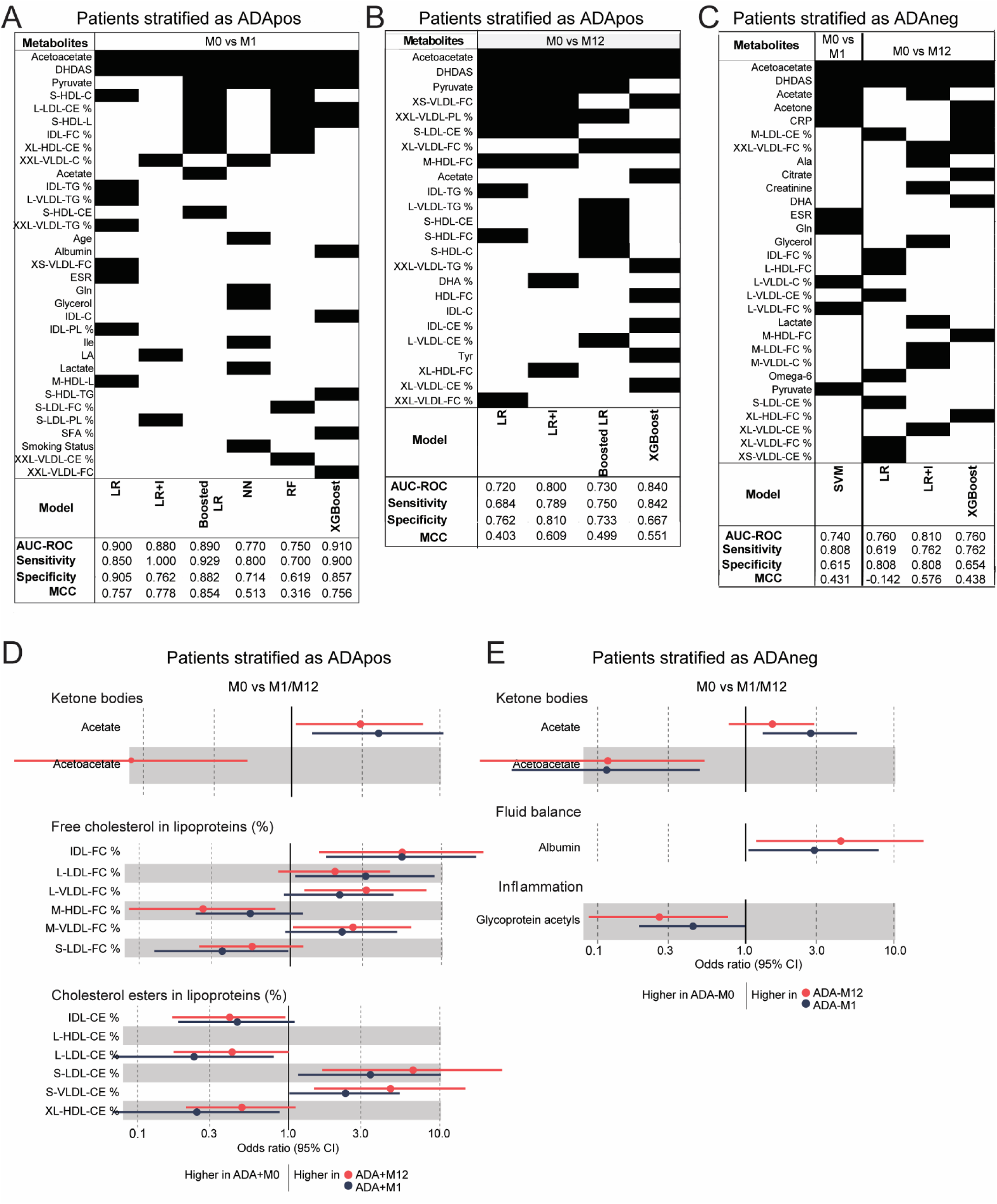
Altered serum metabolite trajectories in RA patients stratified by ADA status. Patients with RA prior to first treatment with adalimumab were stratified based on anti-drug antibody (ADA) status at M12 post-treatment initiation (ADApos, n = 21; ADAneg, n = 26). **(A–C)** Clinical data and serum metabolites were analysed using multiple machine learning models (see Methods). The top features identified by models with AUC-ROC > 0.7 are summarized. **(A)** ADApos patients: M0 vs. M1; **(B)** ADApos patients: M0 vs. M12; **(C)** ADAneg patients: M0 vs. M1 and M12.**(D–E)** Univariate logistic regression forest plots showing the odds ratios (OR) and 95% confidence intervals (CI) for metabolite associations with ADA-positivity **(D)** and ADA-negativity **(E).** Each point represents the OR for a given variable, with horizontal lines indicating the 95% CI. Only significant associations are shown.

Metabolites uniquely regulated in ADApos patients, included pyruvate, large and extra-large VLDL-TG, IDL-TG, small HDL-cholesterol esters and HDL-cholesterol, extra small VLDL-free cholesterol and IDL-cholesterol in both M0 vs M1 and M0 vs M12 comparisons (**Figure 3A-B, Supplementary Fig S2F**). In addition, 26 lipoprotein and fatty acid metabolites, three clinical features (age, smoking status and ESR) and six other metabolites were differentially regulated between M0 and M1, while fewer metabolites were regulated in the M0 vs M12 comparison. These findings were consistent with the univariate LR analysis which showed that free cholesterol (FC) and cholesterol ester (CE) content in lipoproteins varied significantly over the sampling period in ADApos patients (**Figure 3D**). Thus, the results suggest that early changes in cholesterol composition in lipoprotein particles could be associated with future ADA development.

In contrast, fewer metabolites were regulated in ADAneg patients overall and only acetone and CRP featured uniquely in both M0 vs M1 and M0 vs M12 comparisons. Of note both CRP and GlycA (markers of inflammation) were significantly regulated in ADAneg patients over time, reflecting that patients were responding to treatment (**Figure 3C, Supplementary Fig S2E,G**).

### Distinctive metabolic differences at baseline associated with ADA development

Since unique metabolic signatures could be identified in patients stratified according to their future ADA status both at baseline (M0) and following adalimumab treatment we assessed whether we could generate a predictive signature for ADA development in RA. Metabolites associated with ADApos and ADAneg status were assessed for their significance in predicting ADA predisposition (**Figure 4A**). Notably, L-LDL-CE% emerged as the most important metabolite; it was the only one that showed a statistically significant difference between ADApos and ADAneg patients at baseline and was consistently identified across multiple machine learning models.

**Figure 4:**
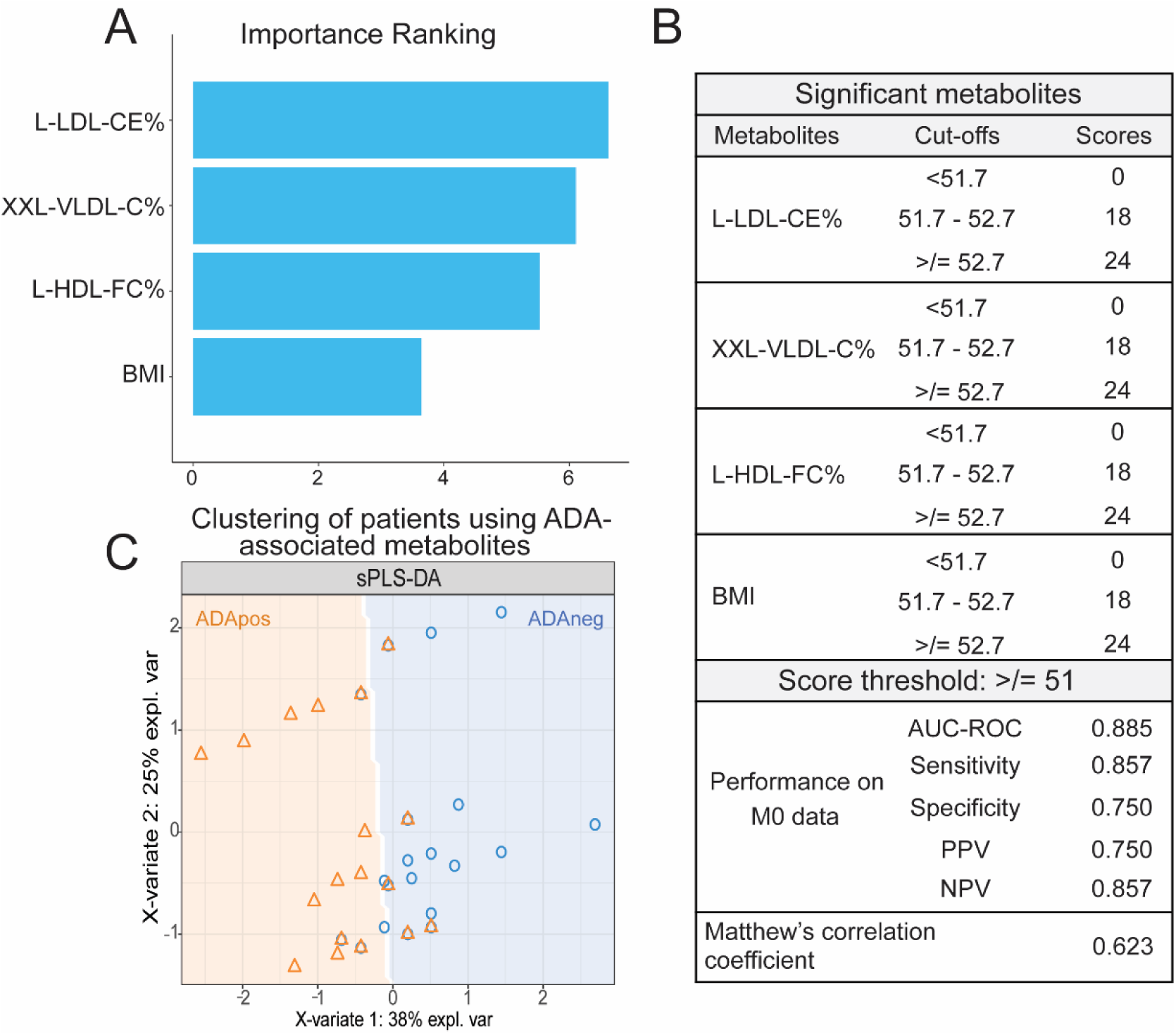
Evaluation of a model generated using ADA-associated metabolites. **(A)** Importance ranking of metabolites included in the final model shortlist. **(B)** The scoring model and its performance in stratifying ADApos and ADAneg patients based on baseline (M0) metabolite levels. (**C)** Clustering of patients based on the metabolite signature using a sparse partial least squares discriminant analysis (sPLS-DA) model. See Supplementary Figure S3 for the ROC curve and loadings for components 1 and 2.

To generate a predictive score, AutoScore was applied, beginning with feature ranking based on associations with ADA status. The highest-ranked metabolites/clinical features were L-LDL-CE%, XXL-VLDL-C%, L-HDL-FC% and BMI, these features were incorporated into a point-based scoring system, where integer-based scores were assigned according to predictive contributions. As previously defined, a well-performing model has a sensitivity and specificity >0.6 and AUC-ROC >0.7. The model was refined to optimise classification performance, achieving a high AUC-ROC (0.885), sensitivity (0.857) and specificity (0.750); these measures suggest that the signature was capable of predicting ADA development with a high degree of accuracy (**Figure 4B**). To assess for overfitting (due to small sample size), Matthew’s correlation coefficient was calculated for the baseline metabolite data, yielding a value of 0.623, indicating a robust model. Markers from the ADA predictive score were also able to cluster ADApos from ADAneg and the combined loadings from components 1 and 2 stratified patients with an AUC-ROC=0.882 (**Figure 4C, Supplementary Figure S3A-B**).

## Discussion

While ML has been used in RA to enhance diagnostic and treatment strategies, its application to ADA prediction remains limited. ML models that integrate omics data, clinical markers, and demographic factors could improve treatment personalization and patient outcomes (29, 30). By uncovering patterns in large datasets, these models can improve understanding of treatment efficacy and optimize RA management (29). Serum metabolites are valuable biomarkers in RA, offering diagnostic and prognostic insights (31, 32). Their clinical appeal lies in the simplicity and minimal invasiveness of blood draws, prompting further exploration in RA care. This study used ML and analytical techniques to identify a small subset of serum metabolites that could predict ADA development in RA patients and identified key metabolite classes that potentially illuminate the mechanisms of ADA development in RA.

At baseline prior to first adalimumab treatment, metabolites related to cholesterol transport and patient-related features including BMI, ESR, smoking status, and sex were able to classify patients based on their future M12 ADA status. Dyslipidaemia, characterised by reduced LDL-C and HLH-C compared to healthy levels is common in patients with active RA (33–35), and is likely due to the effect of proinflammatory cytokines IL-6 and TNF-α, which are known to decrease serum lipid levels (33). Conversely, treatment with TNF inhibitors leads to increased HDL and total cholesterol levels (36). In this study, several lipid metabolites were important for classifying future ADA status in patients with RA suggesting that subtle changes in lipoprotein subsets and composition could contribute to variations in treatment response and play a role in ADA development in RA (37, 38).

BMI was another factor important for classifying future ADA status in the ML models. It is known that high BMI is associated with poor response to TNF-inhibitor therapy in patients with RA(39), which could in part be associated with increased immunogenicity (9). A recent study that also employed ML, showed that low BMI was associated with high drug retention in patients with patients with RA treated abatacept (40). Furthermore in patients with axial spondyloarthritis treated with TNF-inhibitors, high BMI correlated with low drug trough concentration (accelerated drug clearance potentially associated with immunogenicity) (41) and BMI negatively correlated with drug clearance in Crohn’s patients treated with infliximab (42). However, in a study of immunogenicity to infliximab in healthy donors, no significant effect of BMI on drug levels during a 10 week study was seen (43). Elevated-BMI is also associated with elevated CRP, a marker of disease activity in RA, patients with RA having very high or low BMIs tend to have elevated CRP levels(44). Therefore, the association between BMI, cholesterol metabolism and immunity cannot be ignored in the context of future ADA development.

The development of ADAs in RA patients within the first 3-7 months of commencing adalimumab treatment is well recognised (9, 19, 45–47). Our findings were consistent with these observations with most patients developing ADAs by M1 after treatment initiation. As well as having a unique ‘ADA-associated signature’ prior to first adalimumab administration, patients who go on to develop ADAs also had an altered metabolomic response to adalimumab early in their treatment course. Very few metabolites were significantly different in ADAneg patients across timepoints. Those that did persist (acetoacetate, acetate, acetone) as well as clinical features CRP, and DAS-28 were all directly related to disease progression and were likely related to improvement in disease symptoms due to drug action. It has been recognised previously that reduced serum acetoacetate and increased acetate levels are associated with response to treatment in patients with RA(48). We observed similar changes in these ketone bodies in both ADA-pos and ADA-neg patients over the treatment time course despite TNF⍺-inhibition being less effective in the ADA-pos group. TNF⍺ is involved in reprogramming metabolism, specifically of lipids and glucose (49). Inhibition of TNF⍺ could therefore be reflected in changes in the metabolic profile of treated patients (regardless of ADA status), that favours a metabolic shift towards anaerobic glycolysis over aerobic pathways which help to sustains RA pathogenesis by providing energy for inflammatory processes(50, 51).

On the other hand, in ADApos patients, there was a large flux in metabolite concentrations over the one-year period following treatment commencement. Notably, significant changes in lipoprotein composition were dominant, supporting a role for altered cholesterol transport and lipid metabolism in ADA development, as suggested in the ADA-predictive signature prior to treatment. Univariate analysis further underlined the importance of cholesterol content of lipoproteins to ADA development, with VLDL and LDL cholesterol subsets showing many significant differences. VLDLs and LDLs primarily facilitate cholesterol transport to peripheral tissues — a process crucial to maintaining cholesterol balance (52). Increased cholesterol in circulating VLDL/LDLs may indicate elevated delivery to peripheral tissues, potentially increasing cholesterol in lipid rafts, which could influence cellular functions involved in ADA development. Some, like L-LDL-CE and XXL-VLDL-C, were key to ADApos/ADAneg stratification. Circulating lipoprotein cholesterol exists as cholesterol esters (CE) and free cholesterol (FC), either for delivery to or removal from peripheral tissues(52).

Conversely, HDLs are involved in the removal of cholesterol from peripheral tissues. In ADApos patients, FC levels in HDLs were reduced, with L-HDL-FC% being consistently lower at all time periods. This aligns with other RA studies demonstrating that patients with high TGs and low HDL cholesterol respond poorly to TNF⍺-inhibitor treatment (53). Patients with TG^high^HDL^low^ profiles had no reduction in serum TNF⍺, sustaining or worsening symptoms. The authors speculated that this altered profile supports CE exchange, and reduced cholesterol efflux capacity(53). Reduced cholesterol efflux from immune cells can lead to increased cholesterol in the plasma membrane, thereby affecting lipid raft composition and, potentially immune cell signalling(54).

The altered serum lipid profile in ADApos vs ADAneg patients in this study point to potential changes in immune cell cholesterol uptake and efflux via lipoproteins which are known to influence lipid raft composition in immune cell membranes. Lipid rafts – membrane regions enriched with cholesterol, glycophospholipids, and receptors – affect cell signalling and function. Waddington et al. found similar lipid raft changes associated with ADA development in patients with multiple sclerosis, where pre-treatment serum lipids influenced immune cell lipid rafts, increasing ADA risk after the commencement of interferon-beta treatment (16).

The presence of hyperlipidaemia-related pathways aligns with prior studies indicating that dyslipidaemia is not only common in RA patients and can predispose individuals to the disorder(34), but may also contribute to variations in treatment response and immunogenicity(37, 38, 55).

This study highlights several metabolites linked to ADA development, particularly those related to lipid raft composition in immune cell membranes.

Overall, study shows that future ADA development in patients with RA treated with adalimumab could be predicted using a combination of serum metabolomic and clinical/demographic features. Furthermore, the results suggest that alterations in immune-cell lipid rafts may play a crucial role in ADA emergence, warranting further focused research in this area. Certain limitations may affect the interpretation and generalisability of the results, including the small sample size and lack of diversity and these results will need to be validated in a larger study. A metabolite-based approach to predict ADA development in RA patients could support more personalized treatment strategies, allowing clinicians to better anticipate and respond to individual patient needs.

## Data availability statement

All anonymised metabolomic data will be made available on publication

