## Supplementary information for "Serum metabolic signatures are associated with anti-drug antibody development in rheumatoid arthritis patients treated with adalimumab"

**Supplemental Acknowledgments for ABIRISK Consortium**

**List of ABIRISK partners, leaders and task responsible persons**: INSERM (Marc Pallardy, Sophie Tourdot, Xavier Mariette, Sebastien Lacroix-Desmazes, Philippe Broet, Delphine Bachelet, Nadia El-Hamdi); GlaxoSmithKline (Dan Sikkema, Amy Loercher, Julie Davidson, Andy Lawton, Steve Etheridge, Sally Miles); Medizinische Universität Innsbruck (Florian Deisenhammer); UCB Pharma SA (Louis Christodoulou, Hishani Kirby); Academisch Medisch Centrul bij de Universiteit van Amsterdam (Niek De Vries, Anne Musters); Assistance Publique Hopitaux de Paris (Aline Doublet, Mohcine Benbijja); Groupe d'études thérapeutiques des affections inflammatoires du tube digestif (Matthieu Allez, Sabrina Williams); Universitaetsklinikum Bonn (Johannes Oldenburg, Thilo Albert); Heinrich-Heine-University, Düsseldorf (Hans-Peter Hartung, Clemens Warnke, Kathleen Wolffram, Bernd Kieseier); Karolinska Institutet (Anna Fogdell Hahn, Malin Ryner, Ryan Ramanujam); Pfizer (Tim Hickling, Bonnie Rupp); Merck Serono (Elisa Bertotti); Ipsen (Julie Le Grand); University College London (Claudia Mauri, Elizabeth Jury, Jessica Manson); Sanofi-Aventis Research and Development (Vincent Mikol, Agnès Hincelin-Mery, Catherine Prades, Pauline Loas); Università di Firenze (Enrico Maggi); Novartis Pharma AG (Annette Karle, Sebastian Spindeldreher, Verena RomachRiegraf); Fondazione per l'Istituto di Ricerca in Biomedicina (Antonio Lanzavecchia); Klinikum rechts der Isar der Technischen Universitaet Muenchen (Bernhard Hemmer); Commissariat à l'Energie Atomique (Bernard Maillere); Novo Nordisk (Christian Ross Pedersen); Scicross AB (Pierre Dönnes); Bayer Schering Pharma AG (Jeannette Lo, Pascale Buchmann); eTRIKS (Fabien Richard); Paul-Ehrlich-Institut (Christine Keipert); ALTA Ricerca e Sviluppo in Biotecnologie S.r.l.u. (Riccardo Bertini, Simona Farnetani);

**Supplementary Table S1: List of metabolites and their abbreviations.**

| Abbreviation | Full name |
|---|---|
| Total-C | Total cholesterol |
| non-HDL-C | Total cholesterol minus HDL-C |
| Remnant-C | Remnant cholesterol (non-HDL, non-LDL - cholesterol) |
| VLDL-C | VLDL cholesterol |
| Clinical LDL-C | Clinical LDL cholesterol |
| LDL-C | LDL cholesterol |
| HDL-C | HDL cholesterol |
| Total-TG | Total triglycerides |
| VLDL-TG | Triglycerides in VLDL |
| LDL-TG | Triglycerides in LDL |
| HDL-TG | Triglycerides in HDL |
| Total-PL | Total phospholipids in lipoprotein particles |
| VLDL-PL | Phospholipids in VLDL |
| LDL-PL | Phospholipids in LDL |
| HDL-PL | Phospholipids in HDL |
| Total-CE | Total esterified cholesterol |
| VLDL-CE | Cholesteryl esters in VLDL |
| LDL-CE | Cholesteryl esters in LDL |
| HDL-CE | Cholesteryl esters in HDL |
| Total-FC | Total free cholesterol |
| VLDL-FC | Free cholesterol in VLDL |
| LDL-FC | Free cholesterol in LDL |
| HDL-FC | Free cholesterol in HDL |
| Total-L | Total lipids in lipoprotein particles |
| VLDL-L | Total lipids in VLDL |
| LDL-L | Total lipids in LDL |
| HDL-L | Total lipids in HDL |
| Total-P | Total concentration of lipoprotein particles |
| VLDL-P | Concentration of VLDL particles |
| LDL-P | Concentration of LDL particles |
| HDL-P | Concentration of HDL particles |
| VLDL size | Average diameter for VLDL particles |
| LDL size | Average diameter for LDL particles |
| HDL size | Average diameter for HDL particles |
| Phosphoglyc | Phosphoglycerides |
| TG/PG | Ratio of triglycerides to phosphoglycerides |
| Cholines | Total cholines |
| Phosphatidylc | Phosphatidylcholines |
| Sphingomyelins | Sphingomyelins |
| ApoB | Apolipoprotein B |
| ApoA1 | Apolipoprotein A1 |

| Abbreviation | Full name |
|---|---|
| ApoB/ApoA1 | Ratio of apolipoprotein B to apolipoprotein A1 |
| Total-FA | Total fatty acids |
| Unsaturation | Degree of unsaturation |
| Omega-3 | Omega-3 fatty acids |
| Omega-6 | Omega-6 fatty acids |
| PUFA | Polyunsaturated fatty acids |
| MUFA | Monounsaturated fatty acids |
| SFA | Saturated fatty acids |
| LA | Linoleic acid |
| DHA | Docosahexaenoic acid |
| Omega-3 % | Ratio of omega-3 fatty acids to total fatty acids |
| Omega-6 % | Ratio of omega-6 fatty acids to total fatty acids |
| PUFA % | Ratio of polyunsaturated fatty acids to total fatty acids |
| MUFA % | Ratio of monounsaturated fatty acids to total fatty acids |
| SFA % | Ratio of saturated fatty acids to total fatty acids |
| LA % | Ratio of linoleic acid to total fatty acids |
| DHA % | Ratio of docosahexaenoic acid to total fatty acids |
| PUFA/MUFA | Ratio of polyunsaturated fatty acids to monounsaturated fatty acids |
| Omega-6/Omega-3 | Ratio of omega-6 fatty acids to omega-3 fatty acids |
| Ala | Alanine |
| Gln | Glutamine |
| Gly | Glycine |
| His | Histidine |
| Total BCAA | Total concentration of branched-chain amino acids (leucine + isoleucine + valine) |
| Ile | Isoleucine |
| Leu | Leucine |
| Val | Valine |
| Phe | Phenylalanine |
| Tyr | Tyrosine |
| Glucose | Glucose |
| Lactate | Lactate |
| Pyruvate | Pyruvate |
| Citrate | Citrate |
| Glycerol | Glycerol |
| bOHbutyrate | 3-Hydroxybutyrate |
| Acetate | Acetate |
| Acetoacetate | Acetoacetate |
| Acetone | Acetone |
| Creatinine | Creatinine |
| Albumin | Albumin |

| | |
|---|---|
| GlycA | Glycoprotein acetyls |
| XXL-VLDL-P | Concentration of chylomicrons and extremely large VLDL particles |
| XXL-VLDL-L | Total lipids in chylomicrons and extremely large VLDL |
| XXL-VLDL-PL | Phospholipids in chylomicrons and extremely large VLDL |
| XXL-VLDL-C | Cholesterol in chylomicrons and extremely large VLDL |
| XXL-VLDL-CE | Cholesteryl esters in chylomicrons and extremely large VLDL |
| XXL-VLDL-FC | Free cholesterol in chylomicrons and extremely large VLDL |
| XXL-VLDL-TG | Triglycerides in chylomicrons and extremely large VLDL |
| XL-VLDL-P | Concentration of very large VLDL particles |
| XL-VLDL-L | Total lipids in very large VLDL |
| XL-VLDL-PL | Phospholipids in very large VLDL |
| XL-VLDL-C | Cholesterol in very large VLDL |
| XL-VLDL-CE | Cholesteryl esters in very large VLDL |
| XL-VLDL-FC | Free cholesterol in very large VLDL |
| XL-VLDL-TG | Triglycerides in very large VLDL |
| L-VLDL-P | Concentration of large VLDL particles |
| L-VLDL-L | Total lipids in large VLDL |
| L-VLDL-PL | Phospholipids in large VLDL |
| L-VLDL-C | Cholesterol in large VLDL |
| L-VLDL-CE | Cholesteryl esters in large VLDL |
| L-VLDL-FC | Free cholesterol in large VLDL |
| L-VLDL-TG | Triglycerides in large VLDL |
| M-VLDL-P | Concentration of medium VLDL particles |
| M-VLDL-L | Total lipids in medium VLDL |
| M-VLDL-PL | Phospholipids in medium VLDL |
| M-VLDL-C | Cholesterol in medium VLDL |
| M-VLDL-CE | Cholesteryl esters in medium VLDL |
| M-VLDL-FC | Free cholesterol in medium VLDL |
| M-VLDL-TG | Triglycerides in medium VLDL |
| S-VLDL-P | Concentration of small VLDL particles |
| S-VLDL-L | Total lipids in small VLDL |
| S-VLDL-PL | Phospholipids in small VLDL |
| S-VLDL-C | Cholesterol in small VLDL |
| S-VLDL-CE | Cholesteryl esters in small VLDL |
| S-VLDL-FC | Free cholesterol in small VLDL |
| S-VLDL-TG | Triglycerides in small VLDL |
| XS-VLDL-P | Concentration of very small VLDL particles |
| XS-VLDL-L | Total lipids in very small VLDL |
| XS-VLDL-PL | Phospholipids in very small VLDL |
| XS-VLDL-C | Cholesterol in very small VLDL |
| XS-VLDL-CE | Cholesteryl esters in very small VLDL |
| XS-VLDL-FC | Free cholesterol in very small VLDL |

| | |
|---|---|
| XS-VLDL-TG | Triglycerides in very small VLDL |
| IDL-P | Concentration of IDL particles |
| IDL-L | Total lipids in IDL |
| IDL-PL | Phospholipids in IDL |
| IDL-C | Cholesterol in IDL |
| IDL-CE | Cholesteryl esters in IDL |
| IDL-FC | Free cholesterol in IDL |
| IDL-TG | Triglycerides in IDL |
| L-LDL-P | Concentration of large LDL particles |
| L-LDL-L | Total lipids in large LDL |
| L-LDL-PL | Phospholipids in large LDL |
| L-LDL-C | Cholesterol in large LDL |
| L-LDL-CE | Cholesteryl esters in large LDL |
| L-LDL-FC | Free cholesterol in large LDL |
| L-LDL-TG | Triglycerides in large LDL |
| M-LDL-P | Concentration of medium LDL particles |
| M-LDL-L | Total lipids in medium LDL |
| M-LDL-PL | Phospholipids in medium LDL |
| M-LDL-C | Cholesterol in medium LDL |
| M-LDL-CE | Cholesteryl esters in medium LDL |
| M-LDL-FC | Free cholesterol in medium LDL |
| M-LDL-TG | Triglycerides in medium LDL |
| S-LDL-P | Concentration of small LDL particles |
| S-LDL-L | Total lipids in small LDL |
| S-LDL-PL | Phospholipids in small LDL |
| S-LDL-C | Cholesterol in small LDL |
| S-LDL-CE | Cholesteryl esters in small LDL |
| S-LDL-FC | Free cholesterol in small LDL |
| S-LDL-TG | Triglycerides in small LDL |
| XL-HDL-P | Concentration of very large HDL particles |
| XL-HDL-L | Total lipids in very large HDL |
| XL-HDL-PL | Phospholipids in very large HDL |
| XL-HDL-C | Cholesterol in very large HDL |
| XL-HDL-CE | Cholesteryl esters in very large HDL |
| XL-HDL-FC | Free cholesterol in very large HDL |
| XL-HDL-TG | Triglycerides in very large HDL |
| L-HDL-P | Concentration of large HDL particles |
| L-HDL-L | Total lipids in large HDL |
| L-HDL-PL | Phospholipids in large HDL |
| L-HDL-C | Cholesterol in large HDL |
| L-HDL-CE | Cholesteryl esters in large HDL |
| L-HDL-FC | Free cholesterol in large HDL |
| L-HDL-TG | Triglycerides in large HDL |
| M-HDL-P | Concentration of medium HDL particles |

| | |
|---|---|
| M-HDL-L | Total lipids in medium HDL |
| M-HDL-PL | Phospholipids in medium HDL |
| M-HDL-C | Cholesterol in medium HDL |
| M-HDL-CE | Cholesteryl esters in medium HDL |
| M-HDL-FC | Free cholesterol in medium HDL |
| M-HDL-TG | Triglycerides in medium HDL |
| S-HDL-P | Concentration of small HDL particles |
| S-HDL-L | Total lipids in small HDL |
| S-HDL-PL | Phospholipids in small HDL |
| S-HDL-C | Cholesterol in small HDL |
| S-HDL-CE | Cholesteryl esters in small HDL |
| S-HDL-FC | Free cholesterol in small HDL |
| S-HDL-TG | Triglycerides in small HDL |
| XXL-VLDL-PL % | Phospholipids to total lipids ratio in chylomicrons and extremely large VLDL |
| XXL-VLDL-C % | Cholesterol to total lipids ratio in chylomicrons and extremely large VLDL |
| XXL-VLDL-CE % | Cholesteryl esters to total lipids ratio in chylomicrons and extremely large VLDL |
| XXL-VLDL-FC % | Free cholesterol to total lipids ratio in chylomicrons and extremely large VLDL |
| XXL-VLDL-TG % | Triglycerides to total lipids ratio in chylomicrons and extremely large VLDL |
| XL-VLDL-PL % | Phospholipids to total lipids ratio in very large VLDL |
| XL-VLDL-C % | Cholesterol to total lipids ratio in very large VLDL |
| XL-VLDL-CE % | Cholesteryl esters to total lipids ratio in very large VLDL |
| XL-VLDL-FC % | Free cholesterol to total lipids ratio in very large VLDL |
| XL-VLDL-TG % | Triglycerides to total lipids ratio in very large VLDL |
| L-VLDL-PL % | Phospholipids to total lipids ratio in large VLDL |
| L-VLDL-C % | Cholesterol to total lipids ratio in large VLDL |
| L-VLDL-CE % | Cholesteryl esters to total lipids ratio in large VLDL |
| L-VLDL-FC % | Free cholesterol to total lipids ratio in large VLDL |
| L-VLDL-TG % | Triglycerides to total lipids ratio in large VLDL |
| M-VLDL-PL % | Phospholipids to total lipids ratio in medium VLDL |
| M-VLDL-C % | Cholesterol to total lipids ratio in medium VLDL |
| M-VLDL-CE % | Cholesteryl esters to total lipids ratio in medium VLDL |
| M-VLDL-FC % | Free cholesterol to total lipids ratio in medium VLDL |
| M-VLDL-TG % | Triglycerides to total lipids ratio in medium VLDL |
| S-VLDL-PL % | Phospholipids to total lipids ratio in small VLDL |
| S-VLDL-C % | Cholesterol to total lipids ratio in small VLDL |
| S-VLDL-CE % | Cholesteryl esters to total lipids ratio in small VLDL |
| S-VLDL-FC % | Free cholesterol to total lipids ratio in small VLDL |

| | |
|---|---|
| S-VLDL-TG % | Triglycerides to total lipids ratio in small VLDL |
| XS-VLDL-PL % | Phospholipids to total lipids ratio in very small VLDL |
| XS-VLDL-C % | Cholesterol to total lipids ratio in very small VLDL |
| XS-VLDL-CE % | Cholesteryl esters to total lipids ratio in very small VLDL |
| XS-VLDL-FC % | Free cholesterol to total lipids ratio in very small VLDL |
| XS-VLDL-TG % | Triglycerides to total lipids ratio in very small VLDL |
| IDL-PL % | Phospholipids to total lipids ratio in IDL |
| IDL-C % | Cholesterol to total lipids ratio in IDL |
| IDL-CE % | Cholesteryl esters to total lipids ratio in IDL |
| IDL-FC % | Free cholesterol to total lipids ratio in IDL |
| IDL-TG % | Triglycerides to total lipids ratio in IDL |
| L-LDL-PL % | Phospholipids to total lipids ratio in large LDL |
| L-LDL-C % | Cholesterol to total lipids ratio in large LDL |
| L-LDL-CE % | Cholesteryl esters to total lipids ratio in large LDL |
| L-LDL-FC % | Free cholesterol to total lipids ratio in large LDL |
| L-LDL-TG % | Triglycerides to total lipids ratio in large LDL |
| M-LDL-PL % | Phospholipids to total lipids ratio in medium LDL |
| M-LDL-C % | Cholesterol to total lipids ratio in medium LDL |
| M-LDL-CE % | Cholesteryl esters to total lipids ratio in medium LDL |
| M-LDL-FC % | Free cholesterol to total lipids ratio in medium LDL |
| M-LDL-TG % | Triglycerides to total lipids ratio in medium LDL |
| S-LDL-PL % | Phospholipids to total lipids ratio in small LDL |
| S-LDL-C % | Cholesterol to total lipids ratio in small LDL |
| S-LDL-CE % | Cholesteryl esters to total lipids ratio in small LDL |
| S-LDL-FC % | Free cholesterol to total lipids ratio in small LDL |
| S-LDL-TG % | Triglycerides to total lipids ratio in small LDL |
| XL-HDL-PL % | Phospholipids to total lipids ratio in very large HDL |
| XL-HDL-C % | Cholesterol to total lipids ratio in very large HDL |
| XL-HDL-CE % | Cholesteryl esters to total lipids ratio in very large HDL |
| XL-HDL-FC % | Free cholesterol to total lipids ratio in very large HDL |
| XL-HDL-TG % | Triglycerides to total lipids ratio in very large HDL |
| L-HDL-PL % | Phospholipids to total lipids ratio in large HDL |
| L-HDL-C % | Cholesterol to total lipids ratio in large HDL |
| L-HDL-CE % | Cholesteryl esters to total lipids ratio in large HDL |
| L-HDL-FC % | Free cholesterol to total lipids ratio in large HDL |

| | |
|---|---|
| L-HDL-TG % | Triglycerides to total lipids ratio in large HDL |
| M-HDL-PL % | Phospholipids to total lipids ratio in medium HDL |
| M-HDL-C % | Cholesterol to total lipids ratio in medium HDL |
| M-HDL-CE % | Cholesteryl esters to total lipids ratio in medium HDL |
| M-HDL-FC % | Free cholesterol to total lipids ratio in medium HDL |
| M-HDL-TG % | Triglycerides to total lipids ratio in medium HDL |
| S-HDL-PL % | Phospholipids to total lipids ratio in small HDL |
| S-HDL-C % | Cholesterol to total lipids ratio in small HDL |
| S-HDL-CE % | Cholesteryl esters to total lipids ratio in small HDL |
| S-HDL-FC % | Free cholesterol to total lipids ratio in small HDL |
| S-HDL-TG % | Triglycerides to total lipids ratio in small HDL |
| XXL-VLDL-PL % | Phospholipids to total lipids ratio in chylomicrons and extremely large VLDL |
| XXL-VLDL-C % | Cholesterol to total lipids ratio in chylomicrons and extremely large VLDL |
| XXL-VLDL-CE % | Cholesteryl esters to total lipids ratio in chylomicrons and extremely large VLDL |
| XXL-VLDL-FC % | Free cholesterol to total lipids ratio in chylomicrons and extremely large VLDL |
| XXL-VLDL-TG % | Triglycerides to total lipids ratio in chylomicrons and extremely large VLDL |
| XL-VLDL-PL % | Phospholipids to total lipids ratio in very large VLDL |
| XL-VLDL-C % | Cholesterol to total lipids ratio in very large VLDL |
| XL-VLDL-CE % | Cholesteryl esters to total lipids ratio in very large VLDL |
| XL-VLDL-FC % | Free cholesterol to total lipids ratio in very large VLDL |
| XL-VLDL-TG % | Triglycerides to total lipids ratio in very large VLDL |
| L-VLDL-PL % | Phospholipids to total lipids ratio in large VLDL |
| L-VLDL-C % | Cholesterol to total lipids ratio in large VLDL |
| L-VLDL-CE % | Cholesteryl esters to total lipids ratio in large VLDL |
| L-VLDL-FC % | Free cholesterol to total lipids ratio in large VLDL |
| L-VLDL-TG % | Triglycerides to total lipids ratio in large VLDL |
| M-VLDL-PL % | Phospholipids to total lipids ratio in medium VLDL |
| M-VLDL-C % | Cholesterol to total lipids ratio in medium VLDL |
| M-VLDL-CE % | Cholesteryl esters to total lipids ratio in medium VLDL |
| M-VLDL-FC % | Free cholesterol to total lipids ratio in medium VLDL |
| M-VLDL-TG % | Triglycerides to total lipids ratio in medium VLDL |
| S-VLDL-PL % | Phospholipids to total lipids ratio in small VLDL |
| S-VLDL-C % | Cholesterol to total lipids ratio in small VLDL |
| S-VLDL-CE % | Cholesteryl esters to total lipids ratio in small VLDL |

| | |
|---|---|
| S-VLDL-FC % | Free cholesterol to total lipids ratio in small VLDL |
| S-VLDL-TG % | Triglycerides to total lipids ratio in small VLDL |
| XS-VLDL-PL % | Phospholipids to total lipids ratio in very small VLDL |
| XS-VLDL-C % | Cholesterol to total lipids ratio in very small VLDL |
| XS-VLDL-CE % | Cholesteryl esters to total lipids ratio in very small VLDL |
| XS-VLDL-FC % | Free cholesterol to total lipids ratio in very small VLDL |
| XS-VLDL-TG % | Triglycerides to total lipids ratio in very small VLDL |
| IDL-PL % | Phospholipids to total lipids ratio in IDL |
| IDL-C % | Cholesterol to total lipids ratio in IDL |
| IDL-CE % | Cholesteryl esters to total lipids ratio in IDL |
| IDL-FC % | Free cholesterol to total lipids ratio in IDL |
| IDL-TG % | Triglycerides to total lipids ratio in IDL |
| L-LDL-PL % | Phospholipids to total lipids ratio in large LDL |
| L-LDL-C % | Cholesterol to total lipids ratio in large LDL |
| L-LDL-CE % | Cholesteryl esters to total lipids ratio in large LDL |
| L-LDL-FC % | Free cholesterol to total lipids ratio in large LDL |
| L-LDL-TG % | Triglycerides to total lipids ratio in large LDL |
| M-LDL-PL % | Phospholipids to total lipids ratio in medium LDL |
| M-LDL-C % | Cholesterol to total lipids ratio in medium LDL |
| M-LDL-CE % | Cholesteryl esters to total lipids ratio in medium LDL |
| M-LDL-FC % | Free cholesterol to total lipids ratio in medium LDL |
| M-LDL-TG % | Triglycerides to total lipids ratio in medium LDL |
| S-LDL-PL % | Phospholipids to total lipids ratio in small LDL |
| S-LDL-C % | Cholesterol to total lipids ratio in small LDL |
| S-LDL-CE % | Cholesteryl esters to total lipids ratio in small LDL |
| S-LDL-FC % | Free cholesterol to total lipids ratio in small LDL |
| S-LDL-TG % | Triglycerides to total lipids ratio in small LDL |
| XL-HDL-PL % | Phospholipids to total lipids ratio in very large HDL |
| XL-HDL-C % | Cholesterol to total lipids ratio in very large HDL |
| XL-HDL-CE % | Cholesteryl esters to total lipids ratio in very large HDL |
| XL-HDL-FC % | Free cholesterol to total lipids ratio in very large HDL |
| XL-HDL-TG % | Triglycerides to total lipids ratio in very large HDL |
| L-HDL-PL % | Phospholipids to total lipids ratio in large HDL |
| L-HDL-C % | Cholesterol to total lipids ratio in large HDL |
| L-HDL-CE % | Cholesteryl esters to total lipids ratio in large HDL |

| | |
|---|---|
| L-HDL-FC % | Free cholesterol to total lipids ratio in large HDL |
| L-HDL-TG % | Triglycerides to total lipids ratio in large HDL |
| M-HDL-PL % | Phospholipids to total lipids ratio in medium HDL |
| M-HDL-C % | Cholesterol to total lipids ratio in medium HDL |
| M-HDL-CE % | Cholesteryl esters to total lipids ratio in medium HDL |
| M-HDL-FC % | Free cholesterol to total lipids ratio in medium HDL |

| | |
|---|---|
| M-HDL-TG % | Triglycerides to total lipids ratio in medium HDL |
| S-HDL-PL % | Phospholipids to total lipids ratio in small HDL |
| S-HDL-C % | Cholesterol to total lipids ratio in small HDL |
| S-HDL-CE % | Cholesteryl esters to total lipids ratio in small HDL |
| S-HDL-FC % | Free cholesterol to total lipids ratio in small HDL |
| S-HDL-TG % | Triglycerides to total lipids ratio in small HDL |

**Supplementary Table S2: Summary of parameters and R packages used for machine learning comparisons.**

| ML parameters | | | posM0 vs negM0 | posM0 vs posM1 | posM0 vs posM12 | negM0 vs negM1 | negM0 vs negM12 |
|---|---|---|---|---|---|---|---|
| **Number of metabolites** | | | 59 * | 120 | 120 | 111 | 112 |
| **ML model** | **Package** | **Parameter** | | | | | |
| **LR** | glmnet | alpha | 1 | 1 | 1 | 1 | 1 |
| | | lambda | 0.0356 | 0.235 | 0.129 | 0.057 | 0.14 |
| **LR+I** | glmnet | alpha | 1 | 1 | 1 | 1 | 1 |
| | | lambda | 0.0287 | 0.2181 | 0.141 | 0.044 | 0.075 |
| **Bagged LR** | bag | B | 100 | 100 | 100 | 100 | 100 |
| | | vars | 31 | 50 | 6 | 38 | 25 |
| **Boosted LR** | LogitBoost | B | 10000 | 1000 | 1000 | 1000 | 1000 |
| | | nIter | 3 | 4 | 6 | 2 | 2 |
| **NN** | nnet | Size | 2 ** | 2 | 2 | 7 | 7 |
| | | Decay | 0.1 | 0.001 | 0.001 | 1.00E-04 | 1.00E-06 |
| **RF** | rf | ntree | 10000 | 1000 | 1000 | 1000 | 1000 |
| | | mtry | 1 | 10 | 5 | 10 | 10 |
| **SVM** | svmRadial | C | 5 | 4 | 1 | 1 | 5 |
| | | sigma | 0.1 | 0.05 | 0.05 | 0.025 | 0.05 |
| **XGBoost** | xgboost | nrounds | 200 | 100 | 100 | 200 | 100 |
| | | max_depth | 25 | 10 | 10 | 15 | 20 |
| | | eta | 0.1 | 0.1 | 0.1 | 0.1 | 0.1 |
| | | gamma | 0 | 0 | 0 | 0 | 0 |
| | | colsample_bytree | 0.5 | 0.5 | 0.6 | 0.5 | 0.7 |
| | | min_child_weight | 1 | 1 | 1 | 1 | 1 |
| | | subsample | 1 | 1 | 1 | 1 | 1 |

* Percentage only, with reduced clinical markers. Metabolites were reduced to only percentage concentration measurements to optimise performance. Clinical markers were also reduced to DAS, CRP, and ESR.

** Added AST to clinical markers

**Supplementary Table S3: Time to ADA detection and ADA concentrations in ADApos patients.** Summary of the time (in months) to first ADApos test for each patient who developed anti-drug antibodies (ADA) during the study. The table also includes the ADA concentration measured at the time of the first positive test.

| Patient Number | Time of first test for ADApos (# months following treatment commencement) | [ADA] ng/ml |
|---|---|---|
| 1 | 1 | 4.742 |
| 2 | 1 | 5.222 |
| 3 | 1 | 24.057 |
| 4 | 0* | 19.816 |
| 5 | 1 | 40.559 |
| 6 | 6 | 22.571 |
| 7 | 0* | 34.339 |
| 8 | 12 | 20.688 |
| 9 | 1 | 25.138 |
| 10 | 1 | 18.533 |
| 11 | 12 | 17.449 |
| 12 | 1 | 40.779 |
| 13 | 1 | 18.971 |
| 14 | 1 | 13.746 |
| 15 | 3 | 6.725 |
| 16 | 1 | 6.051 |
| 17 | 12 | 5.218 |
| 18 | 1 | 41.328 |
| 19 | 1 | 269.002 |
| 20 | 3 | 14.768 |
| 21 | 1 | 16.503 |
| 22 | 1 | 27.217 |
| 23 | 3 | 6.633 |
| 24 | 1 | 10.532 |
| 25 | 1 | 20.335 |

*Categorised as ADA-pos in metabolomics dataset, because relative electrochemiluminescence (RECL) is greater than >2.08 and % inhibition>23,8% at each time point.

**Supplementary Table S4: Using clinical and demographic features to predict future ADA status.** Demographic characteristics and clinical markers were compared between ADA+ and ADA- patients using machine learning models. Sensitivity, specificity and AUC-ROC are shown for the top performing models.

| **ML Algorithm Performance Using Only Clinical and Demographic Features** | | | | |
|---|---|---|---|---|
| | Logistic Regression | Neural Network | Random Forest | XGBoost |
| Sensitivity | 0 | 0.4762 | 0.3333 | 0.5238 |
| Specificity | 1 | 0.8462 | 0.6538 | 0.5769 |
| AUC-ROC | 0.45 | 0.66 | 0.38 | 0.57 |

**Supplementary Figure S1:**

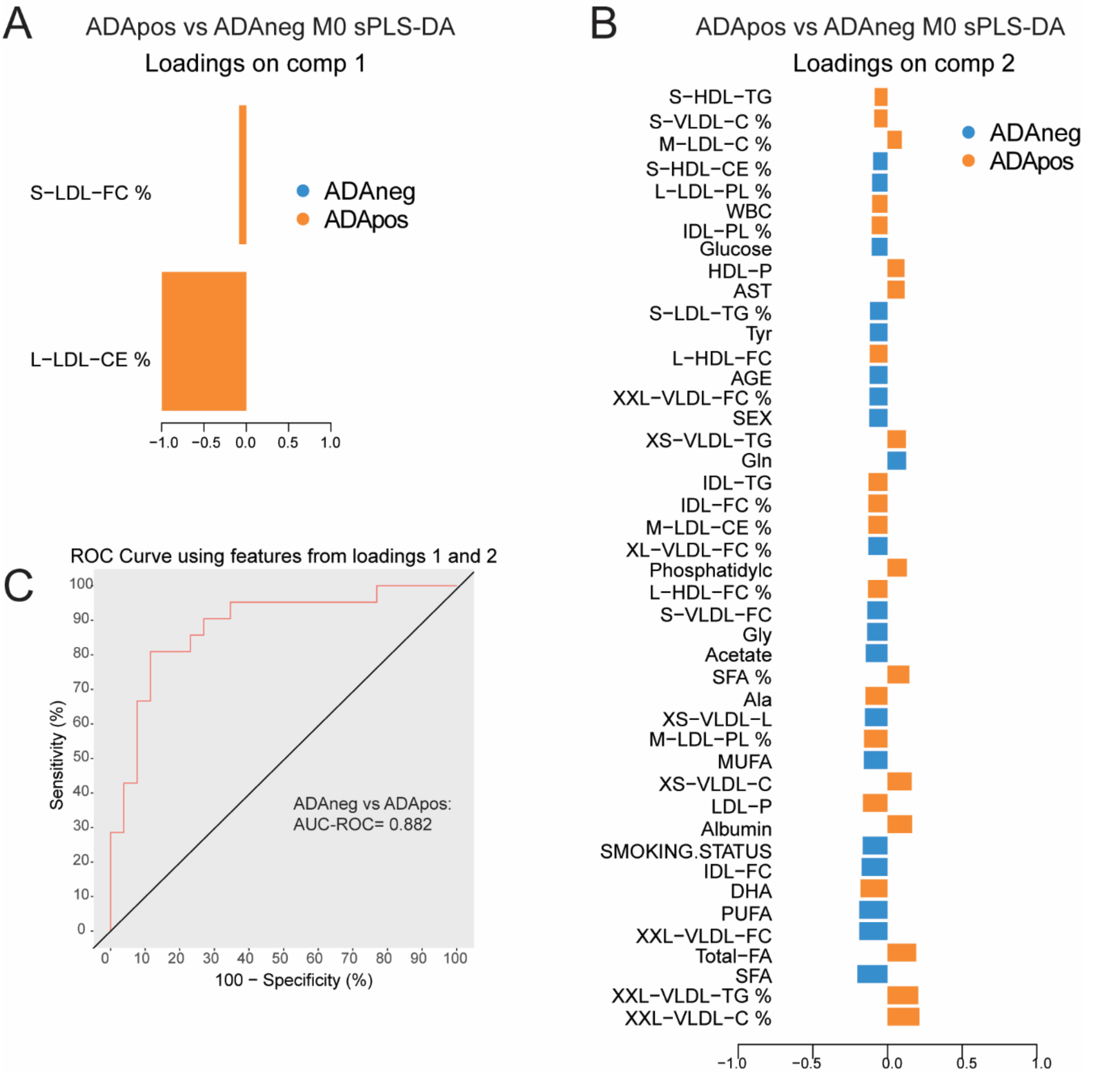


**Supplementary Figure S1: Performance metrics for sparse partial least square discriminant analysis of total metabolites at M0.** Patients with RA prior to first treatment with adalimumab were stratified according to the anti-drug antibody (ADA) status at M12 after treatment initiation (ADApos, n=21; ADAneg, n=26). Data was analysed using a sparse partial least square discriminant analysis model (see Figure 2C). **(A)** Loading of the sPLS-DA shown in Figure 2C for component 1 **(B)** and component 2 are shown. **(C)** The performance of the sPLS-DA in Fig 2C was tested by ROC analysis.

**Supplementary Figure S2**

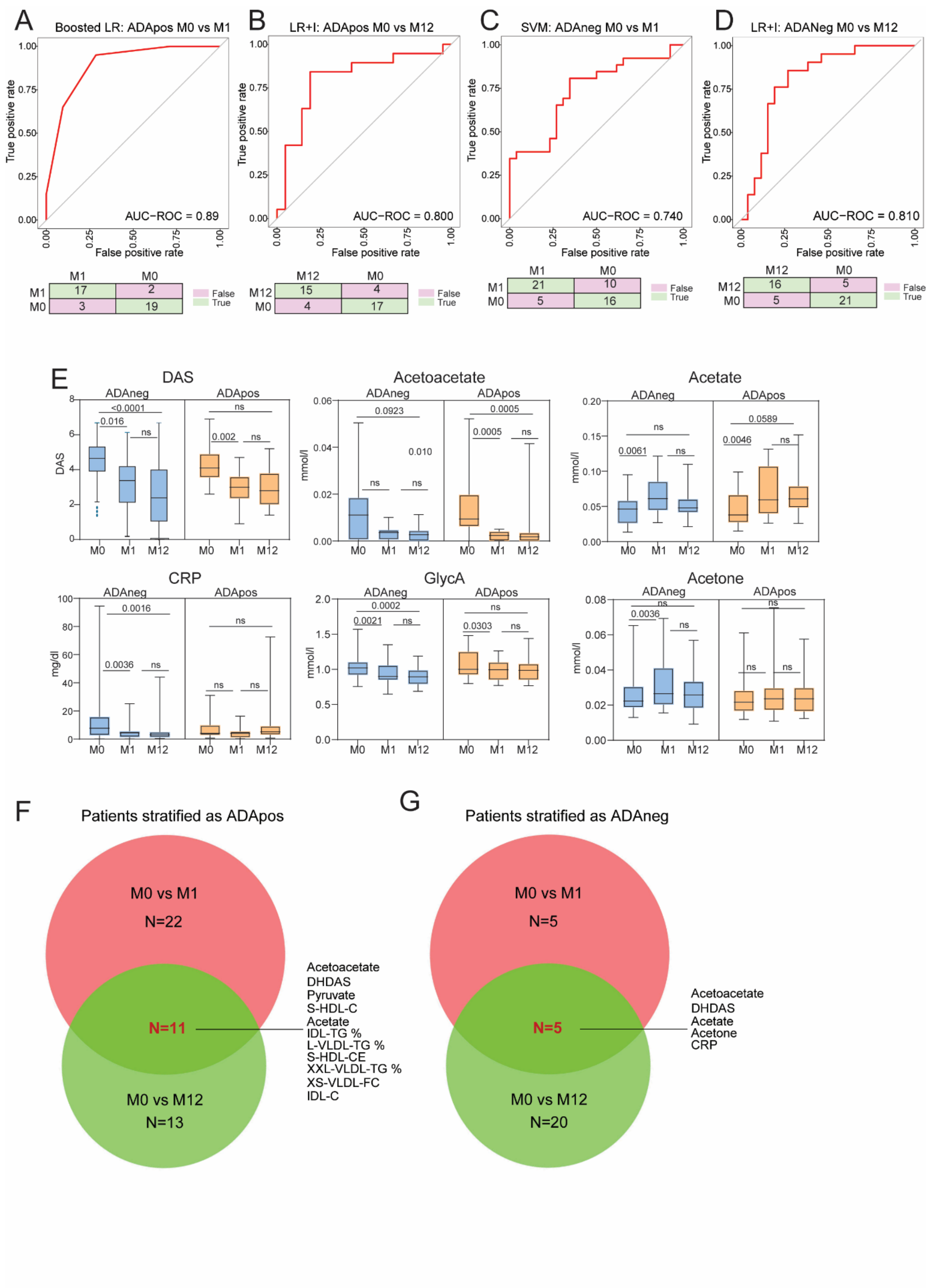


**Supplementary Figure S2: Metabolites combined with clinical features classified patients based on their ADA status determined at M12 following treatment initiation in longitudinal comparisons.** Comparisons are between baseline (M0) metabolite levels and month 1 (M1) and month 12 (M12). Patients with RA prior to first treatment with adalimumab were stratified according to the anti-drug antibody (ADA) status at M12 after treatment initiation. **(A-D)** Receiver operator characteristic (ROC) curve and confusion matrix of the top performing models for each longitudinal comparison in Figure 3A-C. **(E)** Box-and-whisker plots of important features identified by machine learning algorithms. **(F)** Venn diagram illustrating common features between ADAposM0 vs ADAposM1 and ADAposM0 vs ADAposM12 comparisons. **(G)** Venn diagram illustrating common features between ADAnegM0 vs ADAnegM1 and ADAnegM0 vs ADAnegM12 comparisons.

**Supplementary Figure S3**

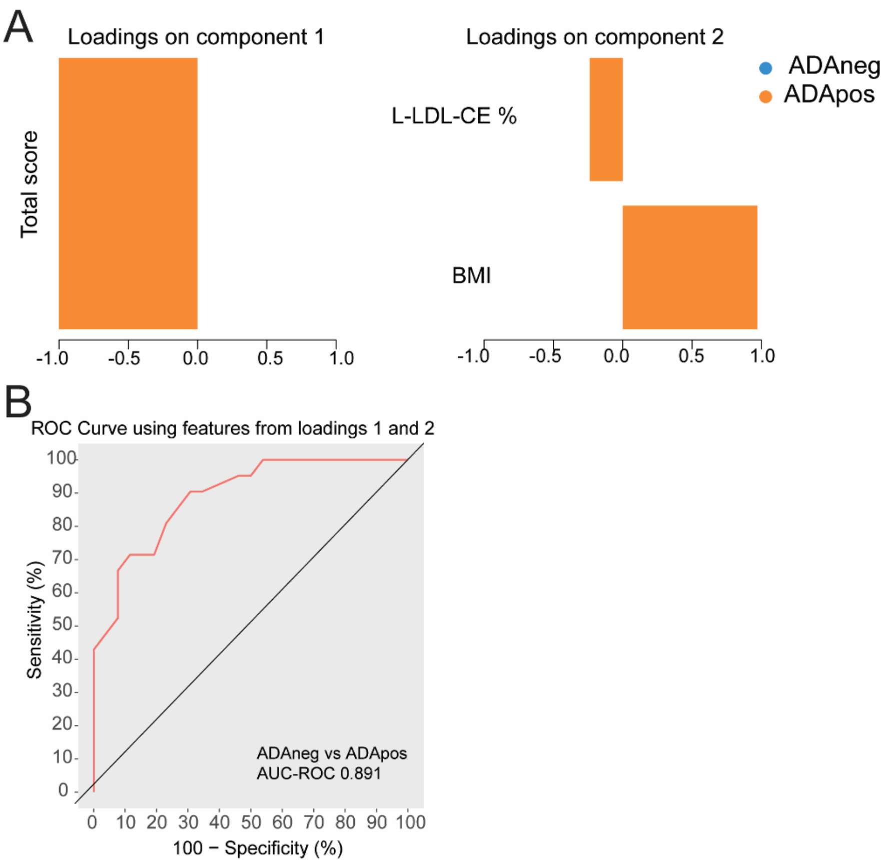


**Supplementary Figure S3: Performance metrics for sparse partial least square discriminant analysis using assigned scores from AutoScore-generated model.** Patients with RA prior to first treatment with adalimumab were stratified according to the anti-drug antibody (ADA) status at M12 after treatment initiation (ADApos, n=21; ADAneg, n=26). Data was analysed using a sparse partial least square discriminant analysis model (see Figure 4C). (**A)** Loading of the sPLS-DA shown in **Figure 4C** for component 1 and component 2 are shown. **(B)** The performance of the sPLS-DA in **Figure 4C** was tested by ROC analysis.
